# Laboratory findings associated with severe illness and mortality among hospitalized individuals with coronavirus disease 2019 in Eastern Massachusetts

**DOI:** 10.1101/2020.05.04.20090555

**Authors:** Victor M. Castro, Thomas H. McCoy, Roy H. Perlis

## Abstract

**Importance:** The coronavirus disease 2019 (COVID-19) pandemic has placed unprecedented stress on health systems across the world, and reliable estimates of risk for adverse hospital outcomes are needed.

**Objective:** To quantify admission laboratory and comorbidity features associated with critical illness and mortality risk across 6 Eastern Massachusetts hospitals.

**Design:** Retrospective cohort study using hospital course, prior diagnoses, and laboratory values.

**Setting:** Emergency department and inpatient settings from 2 academic medical centers and 4 community hospitals.

**Participants:** All individuals with hospital admission and positive severe acute respiratory syndrome coronavirus 2 (SARS-CoV-2) by PCR testing across these 6 hospitals through June 5, 2020.

**Exposure:** Coronavirus 2 (SARS-CoV-2).

**Main Outcome Measures:** Severe illness defined by ICU admission, mechanical ventilation, or death.

**Results:** Among 2,511 hospitalized individuals who tested positive for SARS-CoV-2 (of whom 50.9% were male, 53.9% white, and 27.0% Hispanic, with mean age 62.6 years), 215 (8.6%) were admitted to the ICU, 164 (6.5%) required mechanical ventilation, and 292 (11.6%) died. L1-regression models developed in 3 of these hospitals yielded area under ROC curve (AUC) of 0.807 for severe illness and 0.847 for mortality in the 3 held-out hospitals. In total, 212/292 (78%) of the deaths occurred in the highest-risk mortality quintile.

**Conclusions and Relevance:** In this cohort, specific admission laboratory studies in concert with sociodemographic features and prior diagnosis facilitated risk stratification among individuals hospitalized for COVID-19.

**Funding:** 1R56MH115187-01

**Trial Registration:** None

**Key Points:** *Question:* How well can sociodemographic features, laboratory values, and comorbidities of individuals hospitalized with coronavirus disease 2019 (COVID-19) in Eastern Massachusetts through June 5, 2020 predict severe illness course?

*Findings:* In this cohort study of 2,511 hospitalized individuals positive for severe acute respiratory syndrome coronavirus 2 (SARS-CoV-2) by PCR who were admitted to one of six hospitals, 215 (8.6%) were admitted to the ICU, 164 (6.5%) required mechanical ventilation, and 292 (11.6%) died. In a risk prediction model, 212 (78%) deaths occurred in the top mortality-risk quintile.

*Meaning:* Simple prediction models may assist in risk stratification among hospitalized COVID-19 patients.

## Introduction

With the rapid spread of coronavirus disease 2019 (COVID-19), efforts to predict clinical outcomes and stratify risk have taken on greater urgency as a means of allocating resources and targeting interventions. A recent report of 1099 admitted individuals from China found that 5.0% required intensive care unit (ICU) transfer, and 2.3% required mechanical ventilation^1^. In Lombardy, Italy, around 16% of test-positive individuals required ICU admission^2^. In the United States, characteristics of admitted patients may differ somewhat. A recent case series from the Seattle area described 24 ICU-admitted patients, of whom 75% required mechanical ventilation^3^. In one of the largest U.S. studies to date, among a series of 2,634 hospitalized patients in New York who died or were discharged, 12.2% had required mechanical ventilation^4^.

Given the constrained resources for treatment of COVID-19, particularly with regard to mechanical ventilation, simple approaches to stratifying morbidity and mortality risk at time of hospitalization are needed. In cohorts ranging from 100-200 patients, multiple laboratory studies have been associated with mortality risk, including elevated ferritin, troponin, and C-reactive peptide (CRP)^5^, elevated d-dimer^6^, and low eosinophil count^7^. Most recently, meta-analysis including a total of 3,377 patients identified multiple blood cell indices as most strongly predictive of mortality^8^.

Electronic health records (EHRs) may facilitate rapid and efficient investigation of clinical cohorts, and form the basis of consortia efforts to address COVID-19 at scale^9^. Here, we examined records from 2 academic medical centers and 4 affiliated community hospitals in Eastern Massachusetts. We applied data from 3 of these hospitals to generate simple and transparent models to estimate risk of severe hospital course, characterized by need for mechanical ventilation, intensive care unit level of care, or risk for death, and validated these results in 3 held-out hospitals, including another academic medical center and 2 community hospitals, as a starting point for generalizable efforts at clinical risk stratification^10^. We hypothesized that clinical and laboratory data available at data could efficiently stratify risk for more severe in-hospital course or death.

## Method

### Patients

The full cohort included all individuals age 18 or older hospitalized at any of the 2 academic medical centers and 4 community affiliate hospitals between March 1, 2020 and June 5, 2020, with documented PCR positive test result for severe acute respiratory syndrome coronavirus 2 (SARS-CoV-2) within 5 days of admission. We excluded patients with a severe outcome on the same day of admission (in whom prediction based on laboratory studies would be uninformative) and patients transferred from outside hospitals (CONSORT Diagram). For all of these individuals, prior diagnosis and course during the admission were extracted from the Partners Research Patient Data Registry (RPDR)^11^ and the Enterprise Data Warehouse (EDW) and used to generate an i2b2 datamart^12^. Data were augmented with age, sex, race, and ethnicity from the same source. The enterprise laboratory feed was used to extract SARS-CoV-2 test order and results, as well as additional laboratory values (eTable 1). Laboratory values available in at least 80% of individuals were included in subsequent analysis as continuous measures, after Winsorization at the 99th percentile but otherwise without transformation, along with laboratory-specific high and low flags. As an aggregate measure of comorbidity, Charlson comorbidity index was calculated using coded International Classification of Disease (ICD) 9 and 10 diagnostic codes drawn from the EHR as previously described^13^.

The study protocol was approved by the Partners HealthCare Human Research Committee. No participant contact was required in this study which relied on secondary use of data produced by routine clinical care, allowing waiver of requirement for informed consent as detailed by 45 CFR 46.116. STROBE reporting guidelines for cohort studies were applied.

### Study Design and Analysis

We included all newly-hospitalized individuals with a SARS-CoV-2 positive PCR test within 5 days of admission. Patients were followed from time of admission to hospital discharge or death, with follow-up censored at time of discharge for consistency with similar publications and because mortality data is not yet available from independent sources (i.e., National Death Index or equivalent). The two primary outcomes of interest were 1) COVID severe illness, including any of the following: admission to the ICU, mechanical ventilation or mortality; and 2) mortality. The former composite outcome was selected to avoid the problem of competing risk – i.e., individuals with severe illness who die before either ICU or mechanical ventilation. We selected the earliest laboratory values and vital signs associated with the admission including those measured in the emergency department. Prior ICD diagnoses were grouped using Healthcare Utilization Project (HCUP) Clinical Classification Software (CCS) hierarchy^14^. The log-transformed counts of each CCS diagnosis group were used as predictors. Beyond descriptive analysis, we report appropriate univariate comparisons (i.e., chi square test for binary variables, Student’s t-test for continuous measures) followed by Ll-penalized logistic regression, or the least absolute shrinkage and selection operator (Lasso)^15^, to identify a parsimonious model with sociodemographic features, baseline vital signs, prior diagnosis, and laboratory values as candidate predictors. The hospitals were divided into a training cohort – composed of 1 academic center and 2 community hospital – and evaluative cohort – composed of the other 1 academic medical center and 2 community hospitals. Lasso was applied to all participants with complete laboratory studies in the training cohort, and the performance of the model evaluated in the wholly separate evaluative cohort. Model fitting used all individuals in the training set, with median imputation of missing data (including laboratory values); for individuals in the testing set, we substituted missing values with medians from training set. Model performance was characterized using standard metrics of discrimination and calibration, focusing on the 5 quintiles of risk determined in the training data set, without recalibration.

These logistic regression models offer advantages in interpretability but fail to consider censoring. Therefore, to better characterize model performance in the testing set, for comparison we also utilized survival analysis, right-censoring at time of hospital discharge or end of available data (06/05/2020), presenting Kaplan-Meier curves comparing risk quintile groups. All analyses utilized R 4.0.0^16^.

## Results

The 2,511 individuals hospitalized through June 5, 2020, included 1,348 (53.7%) at academic medical centers and 1,163 (46.3%) at community hospitals; 1,277 (50.9%) were male, 1,354 (53.9%) white, and 679 (27.0%) Hispanic; mean age was 62.6 years (SD: 19.0) (Table 1). In all, 215 (8.6%) were admitted to the ICU, 164 (6.5%) required mechanical ventilation and 292 (11.6%) died. Of the 2,511 total hospitalizations 634 occurred in the testing cohort and 1,877 occurred in the training cohort. Laboratory values are summarized in eTable 2 and illustrated in eFigure 1.

**Table 1.** Sociodemographic characteristics of training and test sets

|  | **Train**<br>**(N=1877)** | **Test**<br>**(N=634)** | **Total**<br>**(N=2511)** | **p value** |
| --- | --- | --- | --- | --- |
| **Hospital type** |  |  |  | 0.150 |
| Academic Medical Centers | 992 (52.9%) | 356 (56.2%) | 1348 (53.7%) |  |
| Community Hospitals | 885 (47.1%) | 278 (43.8%) | 1163 (46.3%) |  |
| **Age at admission, mean (SD)** | 62.1 (19.3) | 64.4 (18.0) | 62.6 (19.0) | 0.007 |
| **Age group** |  |  |  | < 0.001 |
| < 30 | 99 (5.3%) | 22 (3.5%) | 121 (4.8%) |  |
| 30-39 | 189 (10.1%) | 50 (7.9%) | 239 (9.5%) |  |
| 40-49 | 226 (12.0%) | 49 (7.7%) | 275 (11.0%) |  |
| 50-59 | 309 (16.5%) | 111 (17.5%) | 420 (16.7%) |  |
| 60-69 | 316 (16.8%) | 153 (24.1%) | 469 (18.7%) |  |
| 70-79 | 324 (17.3%) | 102 (16.1%) | 426 (17.0%) |  |
| 80+ | 414 (22.1%) | 147 (23.2%) | 561 (22.3%) |  |
| **Male gender** | 983 (52.4%) | 294 (46.4%) | 1277 (50.9%) | 0.009 |
| **Race** |  |  |  | < 0.001 |
| Asian | 70 (3.7%) | 25 (3.9%) | 95 (3.8%) |  |
| Black | 209 (11.1%) | 219 (34.5%) | 428 (17.0%) |  |
| Other | 318 (16.9%) | 71 (11.2%) | 389 (15.5%) |  |
| Unknown | 175 (9.3%) | 70 (11.0%) | 245 (9.8%) |  |
| White | 1105 (58.9%) | 249 (39.3%) | 1354 (53.9%) |  |
| **Hispanic ethnicity** | 563 (30.0%) | 116 (18.3%) | 679 (27.0%) | < 0.001 |
| **Charlson comorbidity index** | 2.559 (3.254) | 2.836 (3.607) | 2.629 (3.348) | 0.072 |
| Mean (SD) |  |  |  |  |
| **ICU admission** | 161 (8.6%) | 54 (8.5%) | 215 (8.6%) | 0.963 |
| **Mechanical ventilation** | 129 (6.9%) | 35 (5.5%) | 164 (6.5%) | 0.234 |
| **Death** | 209 (11.1%) | 83 (13.1%) | 292 (11.6%) | 0.184 |
| **COVID severe outcome** |  |  |  |  |
| (ICU, mechanical ventilation or death) | 338 (18.0%) | 116 (18.3%) | 454 (18.1%) | 0.870 |
| **Discharged to SNF/Rehab** | 253 (40.6%) | 771 (42.1%) | 1024 (41.7%) | 0.506 |

We utilized L1-penalized regression to train a prediction model based on admission characteristics, prior diagnosis and laboratory values in one academic medical center and 2 community hospitals (Table 2). For severe illness, notable features included lymphocytopenia, eosinopenia, neutrophilia, as well as markers of diminished renal function (Table 2). For mortality (Table 2), features were generally similar, with the addition of presence of nucleated red blood cells and other abnormal red blood cell indices, procalcitonin, and greater representation of prior diagnosed codes consistent with pulmonary disease. (Supplemental eTable 3 and 4 report coefficients for features included in penalize regression, without shrinkage).

**Table 2.** Model coefficients

| **Feature** | **Coefficients for COVID Severe Illness Model** | **Coefficients for COVID Mortality Model** |
| --- | --- | --- |
| Age at admission | 0.0129 | 0.0357 |
| Baseline O2Sat | -0.0009 | -0.0107 |
| BUN (mg/dL) (continuous) | 0.0091 | 0.0193 |
| C-reactive protein (mg/L) (continuous) | 0.0014 | - |
| Charlson comorbidity index | 0.0196 | 0.0137 |
| Creatinine (High) | 0.2546 | 0.3607 |
| eGFR (Low) | 0.0736 | 0.0013 |
| EOS (%) (continuous) | -0.3129 | -0.2550 |
| Glucose (mg/dL) (continuous) | 0.0015 | - |
| LDH (U/L) (continuous) | 0.0031 | 0.0014 |
| Lymphocytes (%) (continuous) | -0.0044 | -0.0021 |
| Lymphocytes, absolute (Low) | 0.3049 | 0.1523 |
| MCHC (Low) | - | 0.0099 |
| Monocytes (Low) | 0.2437 | - |
| Neutrophils (%) (continuous) | 0.0007 | - |
| Neutrophils, absolute (High) | 0.2332 | 0.3655 |
| NRBC, absolute (High) | - | 0.3478 |
| Platelets (K/uL) (continuous) | -0.0007 | - |
| Platelets (Low) | 0.1195 | 0.2360 |
| Prior diagnosis of respiratory infections (CCS 8.1) | 0.0804 | - |
| Prior diagnosis of COPD/bronchiectasis (CCS 8.2) | - | 0.0454 |
| Prior diagnosis of dementia/delirium (CCS 5.4) | - | 0.0366 |
| Prior diagnosis of external causes of injury (CCS 18) | - | 0.0238 |
| Prior diagnosis of lung cancer (CCS 2.2) | - | 0.0389 |
| Prior diagnosis of respiratory failure/insufficiency (CCS 8.6) | - | 0.0373 |
| Procalcitonin (ng/mL) (continuous) | - | 0.1823 |
| RDW (%) (continuous) | - | 0.0451 |
| Troponin T (ng/dL) (continuous) | 0.0045 | 0.0030 |
| WBC (High) | - | 0.0064 |

Note: blank entries indicating that the specific feature was not included in the model for the outcome

In the independent testing set composed of a second academic medical center and two other community hospitals, the COVID severe illness model yielded AUC of 0.807 (Figure 1A), with sensitivity of 60.6% and specificity of 88.9% at the top risk quintile (positive predictive value is 54.7%, while negative predictive value is 91.1%). For the mortality model, AUC is 0.847 (Figure 1B); sensitivity of 78.0% and specificity of 87.5% (positive predictive value is 45.6%, while negative predictive value is 96.7%). Both models exhibit substantial lift, with the highest-risk quintile enriched for adverse outcomes in the test cohort (Figure 2A and B).

For illustrative purposes, we also examined COVID severe illness risk quintile (from the model incorporating all adverse outcomes) and mortality risk quintile in Kaplan-Meier survival curves, with curves censored at time of hospital discharge, June 5, 2020 (i.e., end of available follow-up), or 14 days, whichever came first (Figure 3). Quintiles were significantly associated with predicted outcome by log-rank test (X2 = 818, p<0.0001). In total, 212/292 (78%) deaths occurred in the highest-risk mortality quintile.

**Figure 1.**
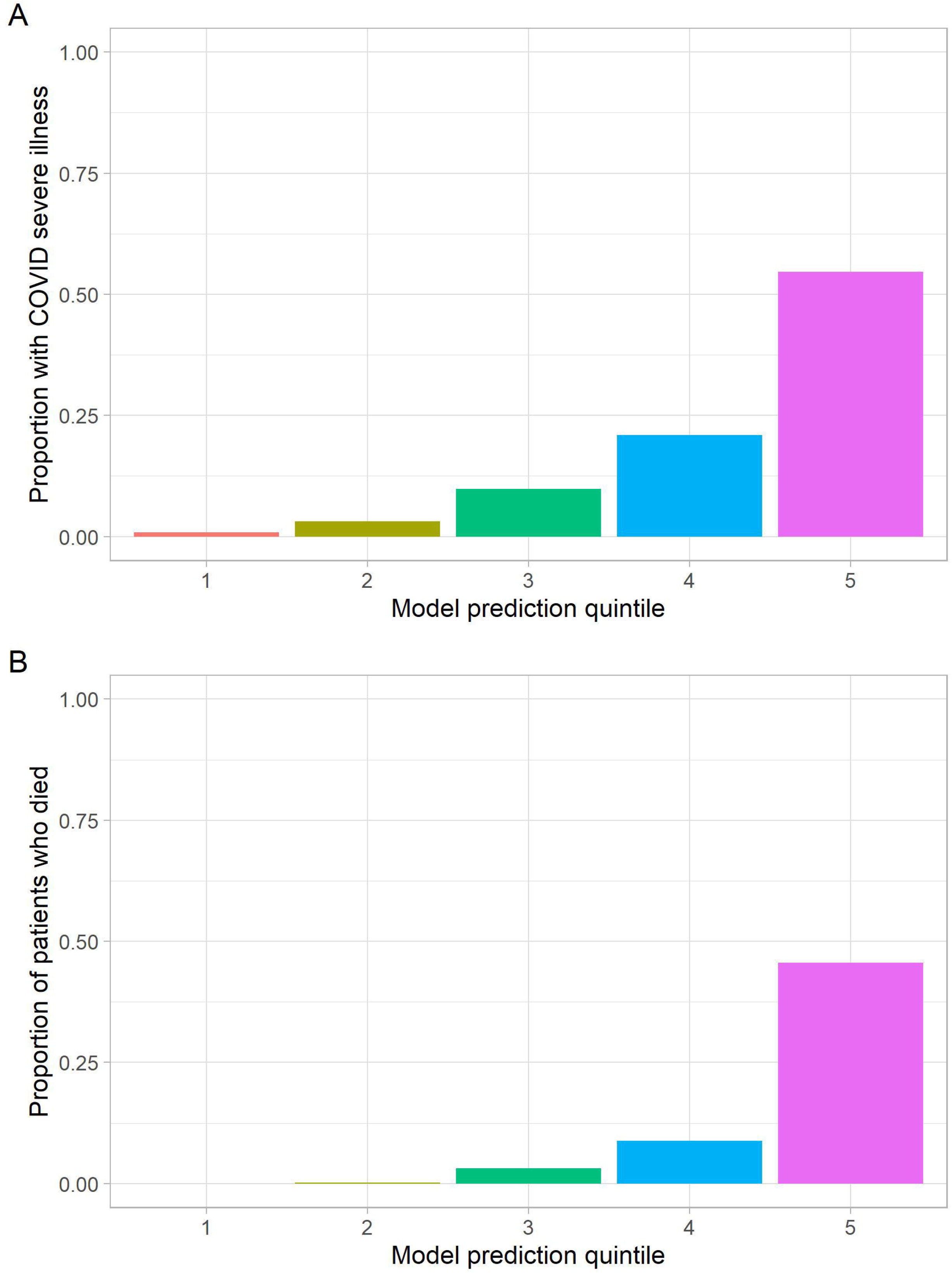
Model Performance in Test Set. A: COVID Severe Outcome B: COVID mortality model

**Figure 2.**
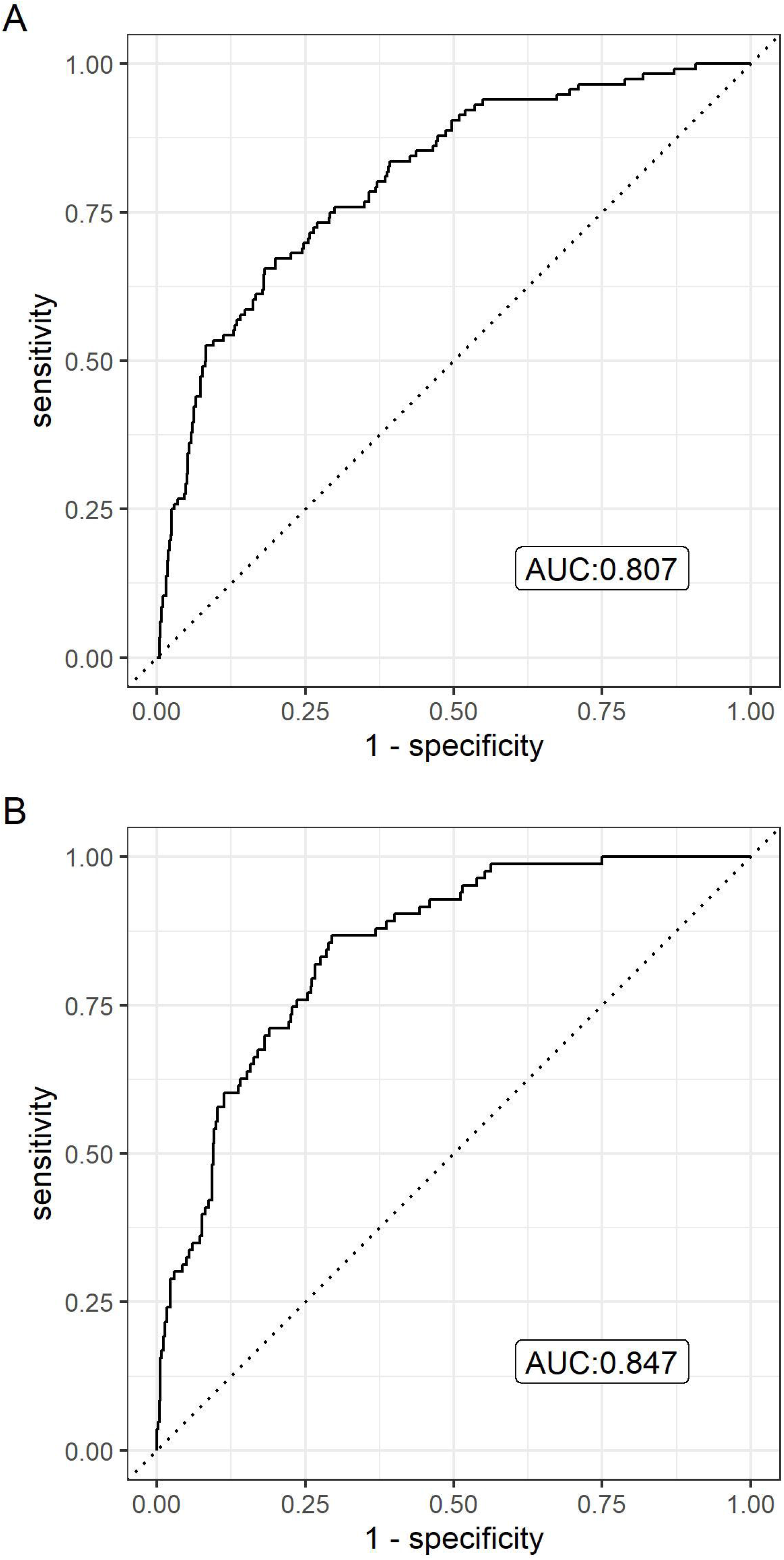
Quintile Plots of outcomes in independent testing cohort

**Figure 3.**
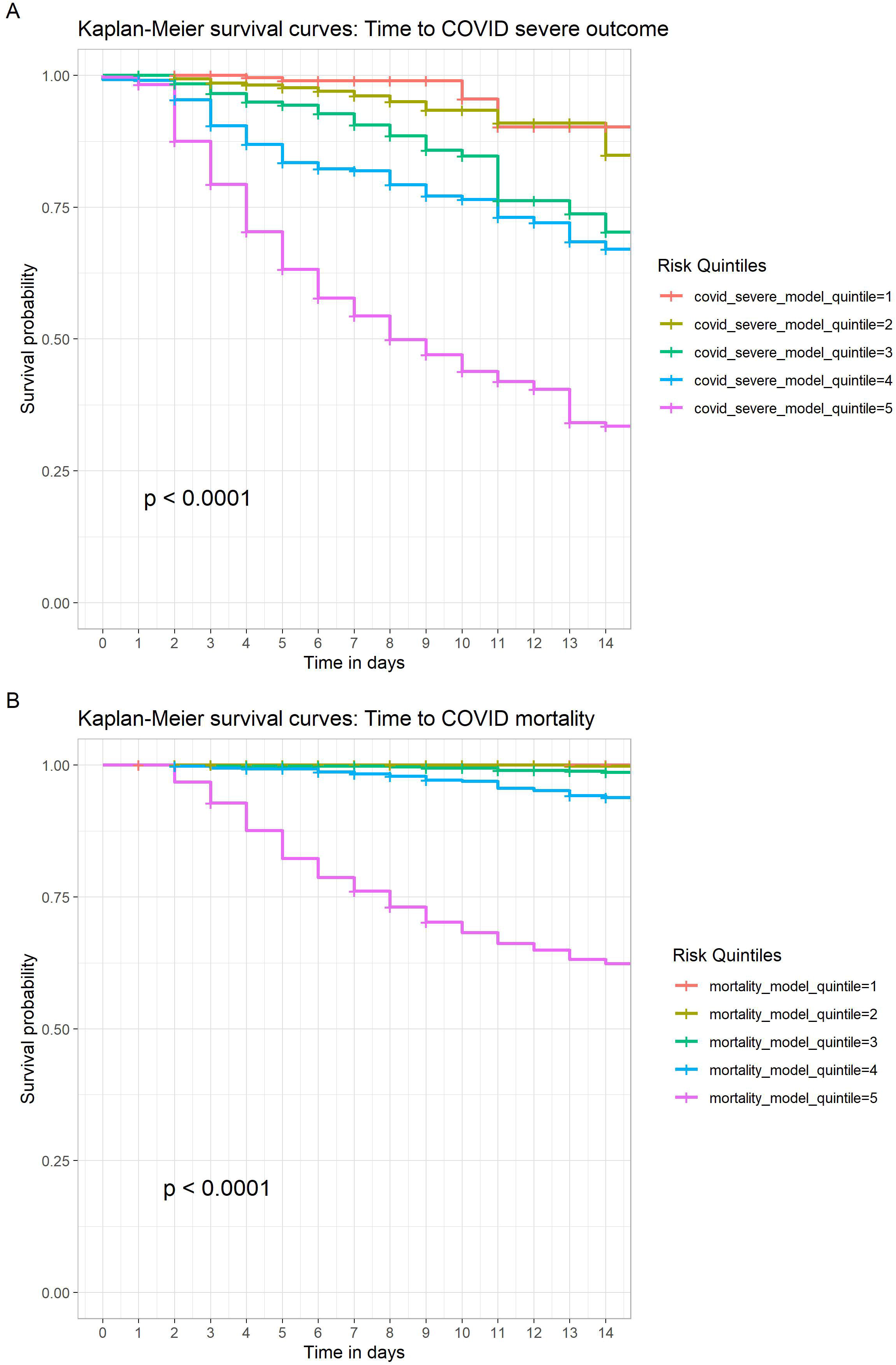
Kaplan-Meier curves in independent testing cohort

## Discussion

In this study of 2,511 individuals with COVID-19 hospitalized at academic medical centers and community hospitals in Eastern Massachusetts through June 5, 2020, 8.6% were admitted to the ICU, 6.5% required mechanical ventilation, and 11.6% died. In general, abnormal hematologic measures (including neutrophilia, lymphocytopenia and eosinopenia), as well as diminished renal function, were associated with greater risk of severe hospital course. Measures of prior pulmonary disease and of red blood cell abnormalities were also represented in risk for mortality.

Discrimination of both models appears promising, identifying a high-risk quintile with reasonable sensitivity and specificity. Likewise, survival curves support the informativeness of the high-risk quintile, and indicate that results are not an artifact of differential attrition or shorter hospital stay. Predictions may be most useful during the initial week of hospitalization; a useful next-step study could examine whether re-running models with additional laboratory studies, or incorporating other biomarkers, can improve longer-term prediction.

Our results are consistent with a recently-reported study associating renal involvement with mortality^17^. Multiple smaller cohorts have also reported laboratory features associated with morbidity and mortality among hospitalized COVID-19 patients. For example, a retrospective cohort study from Wuhan in 191 hospitalized patients found older age and greater d-dimer value at admission were associated with risk of death^6^.

Among 95 fatal cases of COVID-19, low eosinophil count at admission was also common.^7^ Ferritin also associated with mortality in a retrospective cohort study of 120 patients from Wuhan^5^, along with troponin and CRP. Our results also confirm and extend those of the largest meta-analysis to date, encompassing 3,377 patients, implicating hematologic measures as well as renal function^8^, in addition to markers of tissue injury more generally.

In developing these simple prediction models, we were mindful of the recent frameworks for^10^ and criticisms of^18^ such models – particularly the recognition that poorly validated or calibrated models may cause more harm than good. Initial models are likely optimistic (i.e., overfit to data) and biased (i.e., by nonrepresentative samples), with a lack of transparency^18^. On the other hand, strategies to allow risk stratification are particularly necessary in an environment of constrained resources. As such, we report these results in the hope they will provide simple base-case models for others to improve upon. Undoubtedly application of more complex models could yield further improvement in model fit, but whether the degree of improvement is sufficient to offset the added complexity of clinical implementation and reduced interpretability will merit careful consideration.

We note multiple limitations that likely diminish model performance. First, as these are open hospital systems rather than closed networks, lack of documented prior diagnoses does not preclude their presence for individuals who may receive care elsewhere. For this reason we excluded hospital transfers, as prior documentation of comorbidity is likely to be biased. However, such missing data are likely to diminish predictive power of any given diagnosis, such that our model performance estimates are likely to be conservative. In addition, many laboratory values are highly non-normal, such that incorporation of more specific transformations or cut-points could likely improve model performance; we elected to incorporate standard high/low flags plus continuous measures, rather than adopting specific transformations for each value which would risk overfitting or diminish generalizability but likely extract additional information. Efforts to aggregate laboratory data across international health systems will provide an opportunity to explore such transformations if individual-level data becomes accessible^9^.

Despite these limitations, our analyses suggest the utility of laboratory values in combination with documented comorbidities and sociodemographic features in identifying individuals at particularly high risk for more severe hospital course. Notably, by validating in distinct hospitals (albeit within a single geographic region), our estimates of model performance are likely to be less optimistic, but still suggest that generalizability should be good. These admission models also provide an opportunity for comparison as more sophisticated models are developed, particularly those incorporating additional biological measures. To the extent hospital resources are constrained, the ability to target resources to highest-risk individuals is likely to be valuable, and expansion and refinement of risk models may represent a useful approach to optimizing care.

## Data Availability

IRB approval does not allow for sharing of data outside of health system

## Acknowledgements

The authors would like to thank the entire RPDR and Analytics Enclave teams for their support in making up-to-date EHR data available for this research. Dr. Perlis had full access to all the data in the study and takes responsibility for the integrity of the data and the accuracy of the data analysis.

No funding was received for this study. Dr. Perlis has received consulting fees from Burrage Capital, Genomind, RID Ventures, and Takeda. He holds equity in Outermost Therapeutics and Psy Therapeutics. Dr. McCoy has received research funding from the Stanley Center at the Broad Institute, the Brain and Behavior Research Foundation, National Institute of Mental Health, National Human Genome Research Institute Home, and Telefonica Alfa. Mr. Castro reports no conflict of interest.

Dr Perlis had no role in the editorial review or decision to accept the manuscript for publication.

